# Movement guidelines for young children: Engaging stakeholders to design dissemination strategies in the Hong Kong early childhood education context

**DOI:** 10.1101/2022.08.20.22279005

**Authors:** Catherine M. Capio, Rachel A. Jones, Catalina S. M. Ng, Cindy H. P. Sit, Kevin K. H. Chung

## Abstract

**Background:** Early childhood is a critical period during which patterns of movement behaviors are formed. The World Health Organization had endorsed guidelines for physical activity, sedentary behavior and sleep over a 24-hour time period, which had been adopted by the Centre for Health Protection of Hong Kong. This paper reports on stakeholder engagements that were conducted to inform the design of strategies to disseminate the guidelines in early childhood education settings.

**Methods:** Using a mixed-methods study design, we sought to (a) assess the stakeholders’ levels of awareness and knowledge of the Hong Kong movement guidelines for young children, and (b) identify the factors that influence the uptake of the said guidelines. We conducted an online survey of early childhood education teachers (N =314), twelve focus groups involving teachers (N = 18) and parents (N = 18), and individual interviews of key informants (N = 7) and domestic workers who provide care for preschool-aged children (N = 7). Descriptive statistics was used for the quantitative data and thematic analysis was performed on the qualitative data using an inductive and semantic approach following a realist framework.

**Findings:** Our findings show that teachers were aware of the movement guidelines for young children, but their knowledge of the specific guidelines was deficient; parents and domestic workers had limited awareness and knowledge of the guidelines. Uptake of the movement guidelines is enabled by parent engagement, activities in the ECE centers, home-school cooperation, and community activities. The challenges include time poverty of parents, local curriculum requirements, limited physical spaces, social values, and pandemic-related restrictions.

**Conclusion:** We recommend that dissemination strategies in the context of early childhood education settings should deliver knowledge content and support the stakeholders in mitigating the challenges associated with time, space, and social conditions.

## Introduction

The World Health Organization (WHO) released guidelines on physical activity, sedentary behavior and sleep – collectively known as movement behaviors – for children aged five years and younger in 2019 (1). The guidelines were developed as a response to the childhood obesity epidemic and to initiate surveillance and monitoring of children’s movement behaviors over time (2). The Centre for Health Protection (CHP) in Hong Kong adopted the guidelines by the WHO in 2020, but modified them to apply for children aged two to six years, which is the age of children attending pre-primary school (3). This current paper reports on stakeholder engagements that informed the design of dissemination strategies for the guidelines in the context of early childhood education (ECE) in Hong Kong.

Early childhood is widely considered a critical period during which skills and patterns of behaviors are formed, which are believed to track through childhood, adolescence and adulthood (4). In late childhood and adolescence, children tend to have reduced physical activity and increased sedentary time (5,6). The Hong Kong government reports that less than 30% of pre-primary school children (i.e., 3-5 years) accrue 180 minutes of physical activity per day, while the median time spent on sedentary screen time is 60 minutes (7). A longitudinal study showed that 50% of primary school children (i.e., 6-10 years old) met the recommended 60 minutes of moderate to vigorous physical activity per day which declined to 22% two years later (8). It appears that strategies to promote physical activity need to start in the early years, which in the long term does not only contribute to preventing obesity, but also to upholding children’s right to active play (9).

The WHO recognized that whole-day movement behaviors, not only physical activity, contribute to the physical health and well-being of children (10). In earlier work, 24-hour movement guidelines were launched in Canada, Australia, and South Africa, which encompassed physical activity, sedentary behavior, and sleep (11–13). These earlier guidelines informed the WHO recommendations. In Hong Kong, the CHP specified recommendations for two age groups: (a) children aged two years or pre-nursery pupils, and (b) children aged three to six years or nursery, junior kindergarten, and senior kindergarten pupils. There are variations in the recommendations for physical activity and sleep, but those for sedentary behavior are similar for the two age groups (see Table 1 for details). These movement guidelines have been disseminated through ECE centers that are participating in a wider public health campaign on nutrition and physical activity (i.e.). In this current project, we aim to explore and provide suggestions for further dissemination strategies in ECE settings.

**Table 1.**
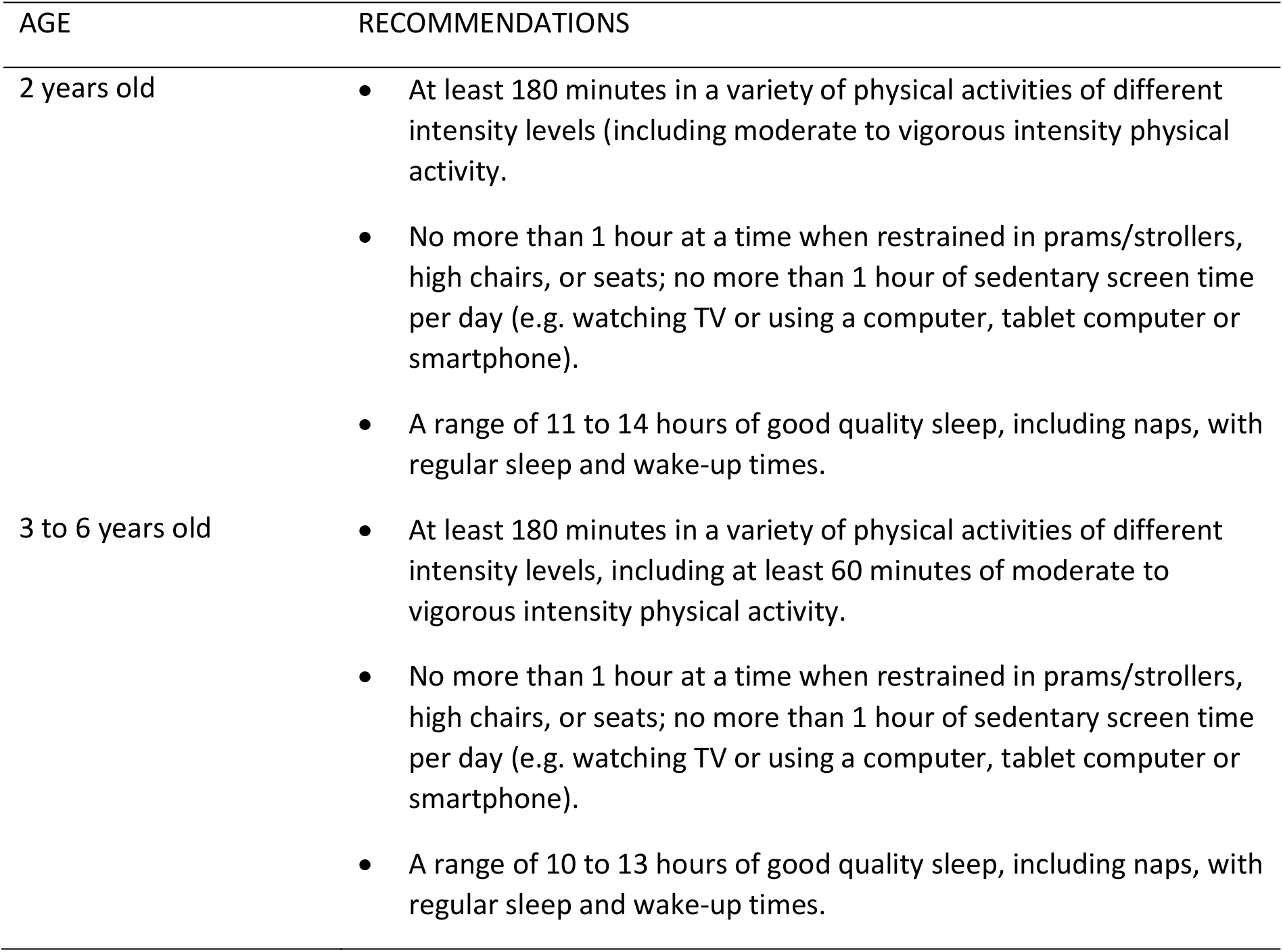
Recommendations for young children in Hong Kong on physical activity, sedentary behavior, and sleep (3).

| AGE | RECOMMENDATIONS |
| --- | --- |
| 2 years old | • At least 180 minutes in a variety of physical activities of different intensity levels (including moderate to vigorous intensity physical activity).<br>• No more than 1 hour at a time when restrained in prams/strollers, high chairs, or seats; no more than 1 hour of sedentary screen time per day (e.g. watching TV or using a computer, tablet computer or smartphone).<br>• A range of 11 to 14 hours of good quality sleep, including naps, with regular sleep and wake-up times. |
| 3 to 6 years old | • At least 180 minutes in a variety of physical activities of different intensity levels, including at least 60 minutes of moderate to vigorous intensity physical activity.<br>• No more than 1 hour at a time when restrained in prams/strollers, high chairs, or seats; no more than 1 hour of sedentary screen time per day (e.g. watching TV or using a computer, tablet computer or smartphone).<br>• A range of 10 to 13 hours of good quality sleep, including naps, with regular sleep and wake-up times. |

To facilitate effective dissemination and uptake of such recommendations, stakeholders’ engagement is essential. Stakeholders consist of expert representatives from the education and health sectors (e.g., ECE administrators, health researchers), practitioners (i.e., teachers, principals), and parents (14,15). The evidence from countries where similar movement guidelines have been launched suggests that stakeholders’ perceptions help assess the acceptability of the recommendations to the local population (16). Qualitative studies have revealed that stakeholders generally found their country-specific movement guidelines acceptable and aligned with health and education priorities (14–16). Barriers to uptake of the guidelines have also been identified, such as lack of available information, daily activities that compete for time and resources, challenges associated with current social norms (14), and high usage of technology-based devices at home (15). The analysis of stakeholders’ needs can also enhance the likelihood of the guidelines’ uptake and identify strategies that can facilitate dissemination and implementation. For instance, it has been suggested that ECE settings provide a natural avenue for information dissemination, where multiple communication channels and a combination of media formats would reach parents and teachers.

While findings from previous research can inform the dissemination strategies of the movement guidelines for young children in Hong Kong, formative context-specific work is imperative. Despite the availability of evidence from other countries, we cannot assume that similar approaches will be suitable and effective. Crucially, health promotion strategies can be successful if local contexts and nuances are analyzed and stakeholders’ needs are addressed (17).

In this current study, we sought to answer the following research questions: (a) *What are stakeholders’ awareness and knowledge levels of the movement guidelines for young children in Hong Kong?* and (b) *What are the factors that influence the uptake of the movement guidelines, which should be considered in designing dissemination strategies?* By answering the first research question, we will be able to assess the starting point for the end users of the guidelines. The second question, on the other hand, will help us leverage on available strengths and mitigate weaknesses in the current systems. Using a mixed-methods study design, we sought to (a) assess the stakeholders’ levels of awareness and knowledge of the Hong Kong movement guidelines for young children, and (b) obtain the insights of key informants, ECE teachers, and parents of children aged two to six years on the factors that influence the guidelines’ uptake. In addition, our initial engagements with the key informants and stakeholders indicated that we needed to engage with domestic workers who contribute to the dynamics of caring for young children in Hong Kong.

## Materials and Methods

Previous studies of stakeholders’ insights have used qualitative designs where interviews revealed individual views, and focus groups explored shared experiences effectively. We adopted a mixed-methods triangulation convergence model to utilize the complementary strengths of quantitative and qualitative methods and gain a comprehensive of the stakeholders’ needs (18). Data were gathered through (a) an online survey, (b) focus group discussions, and (c) individual interviews. All procedures were reviewed and approved by the research ethics committee of the first author’s affiliated university (Reference number 2019-2020-0145).

### Survey

Invitations were sent by email to all registered local ECE centers across the Hong Kong territory. All ECE teachers were eligible to respond to an online survey that was hosted on Qualtrics. Responses were received from 314 teachers, which consisted of 90.0% females as is consistent with the teaching workforce in Hong Kong. The majority of the respondents were aged between 25 to 34 years (42.4%) or 35 to 44 years (30.3%); the rest were aged between 18 to 24 (10.5%), 45 to 54 (11.5%), and 55 to 60 (4.5%) years. Their mean years of experience was 12.4 years (SD = 9.32). Most respondents (58.0%) had a bachelor’s degree qualification; relatively fewer had a sub-degree diploma (28.7%) or a postgraduate degree (13.4%).

The survey consisted of close-ended dichotomous questions (i.e., answered by Yes or No) that were adapted from previous studies to determine the teachers’ awareness (i.e., whether they had seen or heard) and knowledge (i.e., whether they knew the specifications) of physical activity, sedentary behavior, and sleep guidelines (19,20). An open-ended follow-up question asked those participants who claimed to know the guidelines to indicate the minutes or hours recommended for each of the movement behaviors. Further additional questions enquired about the contexts in which the participants promoted the movement guidelines in ECE settings.

### Focus groups

Twelve focus groups were conducted in six ECE centers which involved teachers (n = 18) and parents (n = 18) of children enrolled in the respective centers. One focus group was conducted separately for groups of parents or teachers in each ECE center. The teachers were all females, and they had been in their current roles between two to eight years. The parents were mostly mothers (88.8%). Two experienced researchers facilitated the discussions in the participants’ first language (i.e., Cantonese), which explored the participants’ (a) *awareness and knowledge of the movement guidelines for young children in Hong Kong*, and (b) *perspectives on the factors that impact the uptake of these guidelines*. Open-ended questions were used to initiate the discussions with participants (e.g., “*what makes it difficult for teachers to help reduce sedentary behaviors among pupils?*”). To mitigate the possibility that some participants would dominate the discussion, the facilitators prompted each participant to contribute their insights before transitioning to the next topic (21).

### Interviews

Individual interviews were conducted with the following key informants (n = 7) who were deemed knowledgeable of the local context: ECE principal (for children aged three to six years), nursery program director (for children aged two years and below), education leadership and management consultant, family psychologist, physical fitness specialist, early childhood education academic, and public health academic. The participants had been in their current roles for more than six years, and had been in their respective industries (i.e., education, counselling, health and fitness) for more than ten years.

Two experienced researchers conducted the interviews in English, which explored the participants’ (a) *understanding of the system factors that affect the movement behaviors of young children in Hong Kong*, and (b) *perspectives on how the movement guidelines could be understood and taken up by teachers and parents in the local ECE context*. The open-ended questions were exploratory and sought to obtain participants’ insights based on their own experiences in their respective roles (e.g., “*Based on your experiences with Hong Kong families, what are the factors in the local context that influence the physical activity participation of young children?”, “What do you think are the key considerations if we want parents to be motivated to help their children meet the movement guidelines?*”).

Further individual interviews were conducted with domestic workers who provided childcare for young children (n = 7). In Hong Kong, middle-class families commonly employ a domestic worker as a form of alternative childcare when parents are both working (22). Teachers, parents, and key informants mentioned the roles of domestic workers in children’s movement behaviors. As such, the researchers deemed it of value to seek their insights as well. The domestic worker participants were all females and had been providing care for children in Hong Kong in the last two to six years.

Two experienced researchers conducted the interviews in the participants’ first language (i.e., Tagalog). Open-ended questions explored the participants’ (a) *awareness and knowledge of the movement guidelines for young children*, and (b) *caregiving roles that related to physical activity, sedentary screen time, and sleep of children* (e.g., “*In your daily routines with the child that you are caring for, does he/she engage in physical activities?”, “To what extent are you able to regulate the use of screens/gadgets by the child you are caring for?*”).

### Data analysis

The data gathered from the survey were analyzed using descriptive statistics to determine the percentages of teachers who were aware and knowledgeable of the movement guidelines. The relationships of teachers’ characteristics with their awareness and knowledge of the movement guidelines were examined using Chi-square test (for categorical data) and Spearman’s rank correlation coefficient (for ordinal data). Statistical significance was set at p < 0.05, and all analyses were conducted using IBM SPSS 27.

The data from the focus groups and interviews were transcribed verbatim by researchers who had native proficiency in the language used (i.e., Cantonese, English, Tagalog). The Cantonese and Tagalog transcripts were subsequently translated to English by bilingual researchers. To preserve confidentiality, codes were assigned to each participant according to their group (i.e., T – teacher, P – parent, I – key informant, W – domestic worker) and number (e.g., T01 – teacher, participant 1). These codes are used when quotes are presented in this paper.

A realist framework for qualitative analysis was adopted where language is assumed to capture participants’ experiences of reality (23). Thematic analysis was conducted following a six-phase analytic approach: familiarizing with the data, generating codes, generating initial themes, reviewing and developing themes, refining, defining, and naming themes, and writing the report (24). The research questions of this study (i.e., awareness, knowledge, and factors that influence the uptake of the movement guidelines) directed the focus areas for the analysis. In examining the factors, coding and theme generation were inductive (i.e., bottom up) and semantic (i.e., the explicit meaning of gathered data) to align with the realist framework (23). To ensure the trustworthiness of the analysis, we took a team approach throughout the analytic phases. A team of three members (i.e., the principal investigator and two bilingual researchers each for English/Cantonese and English/Tagalog) read the transcripts independently, where the bilingual members read both the original and translated transcripts to ensure accuracy. Three rounds of iterative discussions were conducted to generate codes and develop themes. The discussions deliberately explored multiple interpretations and reflexivity (25). The interpretations were subsequently discussed with the wider research team as the themes were reviewed and refined.

## Findings

### Stakeholders’ awareness and knowledge of the movement guidelines

The findings from the online survey are summarized in Table 2. The majority of the teachers reported being aware of the guidelines for physical activity (86.9%), sedentary screen time (70.1%), and sleep (58.3%). Most teachers also reported that they knew the recommended minimum time for physical activity (85.4%), the maximum time for using screens/gadgets (79.9%), and the minimum amount of sleep (83.8%) for young children. Based on the open-ended question, it was determined that smaller portions of the teachers reported the correct recommended time for physical activity (19.4%), sedentary behavior (18.2%), and sleep (36.6%).

**Table 2.** Percentage of teachers (N = 314) who are aware and knowledgeable of the movement guidelines.

|  | Movement Guidelines |  |  |
| --- | --- | --- | --- |
|  | Physical Activity | Sedentary Behavior | Sleep |
| Awareness of the guidelines |  |  |  |
| Aware | 86.9 | 70.1 | 58.3 |
| Not sure | 9.9 | 23.6 | 29.6 |
| Not aware | 3.2 | 6.3 | 12.1 |
| Reported knowledge <sup>a</sup> |  |  |  |
| Knowledgeable | 85.4 | 79.9 | 83.8 |
| No idea | 14.6 | 20.1 | 16.2 |
| Actual knowledge <sup>b</sup> |  |  |  |
| Correct knowledge | 19.4 | 18.2 | 36.6 |
| Incorrect knowledge | 65.9 | 61.8 | 47.1 |
| No idea | 14.7 | 20.0 | 16.3 |

*Note:* Data presented as percentage. <sup>a</sup> The respondents reported that they knew the specification of the movement guideline; <sup>b</sup> The respondents who claimed to know of the movement guidelines were asked to indicate the specifications.

Teachers with higher degree qualifications tended to be more aware of the guidelines for physical activity (χ^2^ = 11.11, p = 0.025) and sedentary screen time (χ^2^ = 15.13, p = 0.004) compared to those with lower qualifications. Those who had more years of experience were also more likely to be aware of the guidelines for physical activity (r = 0.171, p = 0.002) and sedentary screen time (r = 0.120, p = 0.033). Neither qualification nor experience was associated with knowledge of the movement guidelines. The frequently reported contexts in which physical activity was promoted in ECE settings included free play periods (86.00%), outdoor activities (77.10%), indoor games (68.80%), and integration with learning areas such as literacy and numeracy (30.90%).

The majority of the teachers who participated in the focus groups reported that they were aware of the movement guidelines (83.33%). All the teachers agreed that the guidelines were suitable for young children in Hong Kong because they were evidence-based. In terms of meeting the guidelines, all the teachers believed that no more than half of their pupils typically meet the daily physical activity guidelines. Furthermore, they believed that most pupils typically exceed the limits for sedentary screen time and do not have adequate sleep.

A small portion of the parents (17.65%) who participated in the focus groups was aware of the movement guidelines for young children. When presented with the specific recommendations for physical activity, sedentary behavior, and sleep, they all agreed that they were suitable for their children. However, the discussion revealed that a minority of them reported that their children can meet the physical activity guidelines (33.33%). There appeared to be relatively more parents who reported that their children can meet the sedentary screen time limits (66.67%) and accumulate adequate sleep (61.11%).

Only one domestic worker was aware and had knowledge of the movement guidelines for young children. While most of them expressed ideas of how much physical activity, sedentary screen time, and sleep are ideal for young children, those ideas did not match with the guidelines. When presented with the correct information of the movement guidelines, none of the domestic workers had an opinion on their suitability for the children. Instead, all of them pointed to the parents as the ones who may judge whether such guidelines are suitable for their children. Only one domestic worker shared routines where the child appeared to meet the movement guidelines; the rest of them described routines where the children appeared to have insufficient physical activity, excessive sedentary screen time, and inadequate sleep. When describing their childcare roles in relation to the movement guidelines, the domestic workers discussed looking after children during outdoor activities and taking them to and from school. About the use of digital devices and sleep, the domestic workers expressed that they generally follow the instructions of the parents.

### Enablers: factors that support uptake of the movement guidelines

#### Teachers

From the teachers’ discussions, two themes were identified as enablers: (a) *parents’ engagement* and (b) *availability of community activities*. The teachers described parents’ engagement in relation to their knowledge and understanding of the movement guidelines, including the impact of physical activity, sedentary behaviors, and sleep on the overall health and development of young children. They further noted that when parents have a good understanding of the *“what”* and *“why”* underlying the guidelines, they are more likely to help their children meet them. Parents’ engagement was also related to their ability to do physical activities with their children at home, regulate the use of digital devices, and implement regular sleep routines. The teachers also believed that parents needed to have a sense of *“how”* to implement the movement guidelines. For instance, one teacher noted: *“For example, tell them (parents) some theory or explanation. Let them know… if the children do physical activity, their development will be affected positively… give them more examples of physical activities – it helps us reach our goal” (T03)*.

The teachers also noted that parenting skills are crucial to establishing routines, managing children’s timetables, and modelling desired behaviors that meet the movement guidelines. Parenting skills were especially discussed in relation to sedentary screen time – e.g. *“Some parents are stricter – they may not really allow children to use devices. However, some cannot control the emotions of the children, so they give the devices when they are acting up” (T15)*.

The teachers recommended workshops to strengthen parents’ understanding of the *“what”* and *“why”* of the movement guidelines. Practical information is needed to provide parents with clear ideas on *“how”* to help children avoid excessive sedentary screen time, and instead accrue adequate physical activity and sleep. For example, a teacher suggested *“let parents know what they can do – recommend them some games to play… find some community resources or exercise video for them to do at home” (T09)*. The availability of activities in the community was discussed in relation to providing practical ideas to parents. Parents could register their children for community activities that are available in their locale (e.g., sports programs, dance lessons) if they were aware that such programs existed.

#### Parents

From the parents’ discussions, two themes were found to be consistent with those shared by the teachers: (a) *availability of community activities* and (b) *support for parents’ engagement*. Similar to the teachers, the parents also expressed the importance of community activities. The parents appeared to rely on community classes when they needed childcare support – e.g., *“They can have community class and someone can take care of them” (P06)*. They also described community activities as means for them to come up with ideas (e.g., *“I can’t think of more activities, so I search for classes” (P05)*), or to plan for family activities (e.g., *“A civic group held parent-child hiking activity and social workers tell us which trails are suitable for kids to hike” (P11)*).

In terms of parents’ engagement, the participants expressed that they could promote healthy movement behaviors if they had support. The parents discussed that they need to have updated knowledge of what is beneficial and what is harmful in relation to physical activities, sedentary behaviors, and sleep. They described some common beliefs that appear to be inaccurate such as children not needing to exercise, or digital screens being harmful to the eyes. It also appeared that parents need to be prompted to seek detailed information about the movement guidelines, as evident here: *“I don’t search for information, I just know exercise and sleeping are important for the growth of children” (P04)*.

The parents also expressed that they could promote physical activity if they had practical ideas. For instance, one parent mentioned: *“What can we do within such a small space at home? Maybe you can provide more activity suggestions with different materials that we can find at home?” (P03)*. Support in relation to sedentary screen time was frequently mentioned and appeared to be crucial as evident in the following examples: *“When I really can’t think of any activities, I only can let him watch the screen” (P06)* and *“…let us know more about family games which can reduce children’s screen time” (P05)*.

Two other enabling factors were identified in the parents’ discussions: (a) *roles played by the ECE teachers*, and (b) *opportunities for children to socialize*. The parents believed that ECE teachers have the knowledge about movement behaviors and are integrate them into their teaching. For example, one participant highlighted that teachers are more knowledgeable than parents: *“In fact we lack systematic knowledge to implement WHO standards… schools can systematically include [them] into the syllabus” (P13)*. It was also noted that teachers could explicitly teach their pupils about healthy movement behaviors, which the parents expect to have a greater influence on the children: *“children will listen to the teacher’s word, like if the teacher said that they cannot watch screens for a long time, it is better than parents telling them” (P15)*.

The parents recognized that opportunities to socialize with other children is an important enabler because it promotes consistent engagement in activities. The presence of playmates in outdoor settings (e.g., playground, park) tends to make children stay outside longer. Siblings also appeared to encourage children to participate in physical activities. One parent noted that *“Children persist when there is a group engaging together in the sport” (P10)*, while another parent mentioned that *“I have three kids, they are mates of each other when doing physical activity” (P13)*.

#### Key informants

From the key informants, *support for parents’ engagement* was also identified as an enabling factor. While their responses were consistent with parents’ discussions, the key informants highlighted that besides strengthening parents’ knowledge, facilitating parents’ motivation to promote the recommended movement behaviors is needed. To enhance motivation, supporting programs need to counter parents’ beliefs that physical activities and play are not as important as academic pursuits, as illustrated here: *“Parents need to understand that physical and cognitive development are equally important” (I02)*. Drawing from their experiences with local families, the key informants noted the importance of acknowledging the constraints faced by working class parents. For instance, one key informant mentioned that *“Parents from regular families are already exhausted after work, and they get home around 8pm or later; this is a huge challenge for them to think of regular physical activities for their children” (I01)*. Workshops may help improve parents’ knowledge, but support should be consistent as mentioned here: *“support for parents cannot be one-off… it has to be designed to motivate long-term results” (I05)*.

The key informants identified two other enabling factors: (a) *home-school cooperation*, and (b) *support for ECE teachers in their roles*. Cooperation between the parents and teachers was deemed an essential component of promoting the uptake of the movement guidelines. The key informants from the education sector acknowledged the packed ECE curriculum and teachers consistently try to meet the government-mandated requirements, which tends to limit their capacity to integrate movement behaviors into their curricula. Activities across a continuum from school to home, therefore, are crucial to improve the uptake of movement guidelines. Moreover, the key informants believed that teachers are effective messengers who can share knowledge with parents and encourage changes in mindset and routines. For instance, one participant mentioned that *“Some parents may not have the habit of physical activity themselves, or do not understand the guidelines… schools can help share knowledge” (I03)*.

As noted above, parents also identified the role of ECE teachers in enabling the uptake of the movement guidelines. For the key informants, however, the enabler is the support provided to the teachers so they can deliver the role of enhancing parents’ awareness and knowledge. To this end, one key informant described it as *“Health experts should train teachers, so they would in turn know to manage and help parents in promoting the movement guidelines at home” (I02)*. As ECE teachers need to design classroom activities within the constraints of the curricular demands, it is crucial that principals buy into the value of the movement guidelines because they have the authority to endorse curricular modifications. Thus, one form of support is targeted messaging for principals. The key informants further described support in the form of on-site and bespoke workshops that will enhance the teachers’ ability to design classroom and home-based activities that are integrated into learning areas and daily routines. One key informant mentioned that *“Teachers need to appreciate that it does not have to be physical education… instead, it has to be integrated with lessons and routines” (I06)*.

### Challenges: factors that hinder uptake of the movement guidelines

#### Teachers

The teachers’ discussions revealed four themes as challenges to children meeting the movement guidelines: (a) *curriculum requirements that lead to time constraints*, (b) *limited space*, (c) *time poverty of parents*, and (d) *restrictions due to the COVID-19 pandemic*. The most widely discussed challenge was related to the local curriculum requirements. It was noted that the academic learning areas (e.g., numeracy, writing) tend to fill up the three-hour daily classes. Thus, the teachers find it difficult to find the time to implement physical activities. As parents are aware of the curricular requirements, they also expect homework to be academic-oriented. Thus, time at home tends to be focused on academic homework – *“For the homework, parents want to see their children complete their academic work. So, the playing time in the playground becomes limited” (T02)*. It was also noted that recent changes in local regulations meant there is no more option for nap time unless the children are attending a full-day program.

The teachers discussed that ECE centers mostly have limited space, which presented a challenge to implementing physical activities as mentioned here, *“the physical environment of the center is not big, so activities are limited” (T07)*. They seek options outside the center (e.g., community sports grounds) but they are typically less prioritized – *“it is difficult for ECE schools to book sports grounds; it is easier for primary and secondary schools” (T09)*. As such, activities tend to be mostly indoors, which teachers find challenging in relation to sustaining the children’s interests – e.g., *“If they always play the same things, they may feel bored” (T17)*. The teachers also noted space issues are also present at home; hence, they need to support parents by giving them activities that can be done in small spaces.

With regards the home situations, the teachers noted that parents typically do not have enough time to engage their children to meet the movement guidelines. They had observed that parents’ working hours affect the children’s routines, especially in relation to sleep – children sleep late because parents get home late from work. Moreover, the teachers believed that parents have difficulty regulating screen time because they prefer that their children do not get upset, as shown here – *“If parents take the mobile phone away, the children will be upset. Then, parents will not remove the mobile phone because they need to get out for work” (T04)*. Long working hours also mean that parents tend to be exhausted and have no energy to engage in physical activities with their children, as illustrated here: *“Most parents want to rest, and so don’t want to exercise. As a result, their kids will not exercise too” (T11)*.

Working parents manage the time issues by relying on the grandparents or domestic workers to look after the children, but the teachers noted that such arrangements often lead to children spending more time indoors and being sedentary. For instance, one teacher mentioned that *“Some children say that their grandparents love to let them watch (the) iPad, but when their mum is back they can’t watch anymore” (T08)*. Thus, even if parents were to gain more knowledge about the movement guidelines, implementing them may remain a challenge as parents attempt to manage their time by relying on the grandparents or domestic workers.

The last challenge identified by the teachers relates to the restrictions due to the COVID-19 pandemic. The most frequently mentioned issue was the suspension of schools, and the teachers noted the impact on physical activities. In 2020, schools across all levels in Hong Kong had suspended periods during which time children were mostly confined to their homes. The teachers observed that many parents were concerned of infection and thus kept their children indoors, as evident here – *“parents said that with the pandemic, they refused to bring their kids to the park as they thought it is risky” (T07)*. Moreover, playgrounds and public spaces were closed such that even if parents allowed their children to go out, the spaces were restricted. Finally, despite curricular limitations, the teachers thought that children were able to accrue greater physical activity when they attended school compared to the period of school suspension. As one teacher noted, *“we provide some physical activity time for children and they may ride a bike or walk to school” (T14)*.

#### Parents

The following challenges were identified from the parents’ discussions: (a) *time poverty*, (b) *limited facilities and space*, (c) *restrictions due to the COVID-19 pandemic* and (d) *values held by the Hong Kong society*. The parents confirmed that they struggle with finding time to help their children meet the movement guidelines. This is especially true for those families where both parents are working. For example, they expressed not being available to engage with their children – e.g., *“Most of us are both working parents and there is a problem of who will do activities with the children” (P02)*. Due to limited time, grandparents and domestic workers assume childcare roles, which the parents acknowledged are not conducive to children meeting the movement guidelines. For instance, one parent noted that *“Screen time will be a bit difficult to control as me and my husband need to work, grandparents mainly take care of them (children). My kids are active, the one thing that grandparents can use to calm them down is the phone” (P09)*.

Families where one parent is full-time at home appear to be more capable of promoting the movement guidelines. Parents are able to implement routines and be firm with them, as evident here *“As a full-time mum, I will bring him to the park every day and let him have enough time to have moderate intensity exercise” (P07)*. Nevertheless, home duties also tend to impact on sleep patterns when routines are not set – e.g., *“My kids don’t go to bed so early because they insist on sticking with me while I finish housework” (P11)*.

The parents also discussed the challenges associated with limited spaces at home and in the community. Combined with parents’ time poverty, the typically small homes in Hong Kong mean that parents can promote physical activities only on days that they are off work, as mentioned here *“there is not enough space, but we can’t take them out to play on weekdays” (P07)*. The problem of space also appears to be related to parents’ lack of knowledge as evident here *“we don’t know what to do with them in such a small space at home” (P01)*. The parents appeared to rely on shared community spaces as venues for physical activity, which include public playgrounds, parks, and rooftop areas. However, they deemed these community spaces inadequate and inconsistent – *“other places now have more parks for children but this is not the same for all places; there are more spaces dedicated to shopping malls” (P08)*. Many community facilities also charge fees, which adds another layer of challenge for many working class families as shown here – *“we need to reserve venues and it is expensive to pay $60 per hour” (P14)*.

For the parents, the main issue with the pandemic-related restrictions was that children were not able to go out either to go to school or to play outdoors. Parents discussed that without school activities, their children tended to be inactive as evident here: *“before the pandemic, I mostly rely on the school for exercise” (P17)*. Moreover, the challenges associated with small home spaces were aggravated. Some parents discussed attempts to do structured activities for their children, but these were infrequent and perceived as additional burden. These are evident in the following: *“only a few times, I managed to design an obstacle course at home for him” (P04), “I feel tired thinking of what to do with him every day” (P06)*. Even after the restrictions were lifted and children were able to use community spaces, socialization opportunities did not really resume – *“after the park was re-opened, my children cannot find constant friends so they felt bored” (P13)*.

Finally, the parents identified challenges associated with the values upheld by the Hong Kong society. The most discussed point is the importance of academic achievement, which encourages parents to focus on academic learning beginning from ECE. Despite focusing on academics themselves, most of the parents appear to be aware that it is not helpful with the movement guidelines – *“kids have lots of things to learn like English so they may not have too much time to play” (P06)*. Some parents also noted that physical activity and sports are not generally valued in Hong Kong – *“Hong Kong people don’t like sports” (P11)*, which they felt makes it difficult to find opportunities for their children to join. Social values also contribute to grandparents who are not supportive of letting children engage in physical play.

#### Key informants

Similar to the challenges identified by the teachers and parents, the key informants identified the following: (a) *time poverty*, (b) *curriculum requirements*, and (c) *values held by the Hong Kong society*. The challenge associated with working parents’ limited time came up once again, where key informants from the education sector noted that working parents could not actively promote the movement guidelines even if they had knowledge of them. For example, *“Parents will know the guidelines, but they will need to go to work” (I03)*, and *“Parents are already exhausted with work to follow through with the guidelines” (I01)*.

Key informants from the health and family sector also noted that in the current curriculum, *“there is very little room for ECE to involve physical activity” (I04)*. Consequently, teachers need to focus on meeting academic expectations and eventually have little practice with promoting physical activities – *“Teachers don’t get the opportunities to explore and be better” (I05)*. Finally, the prevailing values of the Hong Kong society were related to high levels of concern for safety. Informants from both the education and health sectors noted that safety is always prioritized by both parents and teachers, limiting the opportunities for physical play – *“Play should be an adventure, but they (parents, teachers) put safety as the major priority (I02)*.

Combining the responses of teachers, parents, and key informants, four enablers and five challenges were identified in relation to promoting the uptake of movement guidelines for young children (see Table 3). These factors may be considered in designing school-based dissemination strategies of the movement guidelines.

**Table 3.** Factors that enable and challenge children’s compliance with movement guidelines.

| Factor | Theme | Description |
| --- | --- | --- |
| Enablers | Parent engagement | This consists of parents' knowledge of the movement guidelines, understanding of the rationale, and knowledge of strategies to implement the guidelines at home. This also involves parenting skills that facilitate children towards meeting the guidelines. Accessible support for parents underpins parent engagement.<br>Parenting skills include parents' policies at home to set routines (e.g., sleeping time) or regulate activities (e.g., screen time) of children. This also relates to the consistency between parents' knowledge and what is implemented at home. |
|  | Role of ECE centers | This relates to the curriculum content in ECE centers and learning activities that enhance physical activity participation, reduce sedentary screen time, and promote healthy sleep patterns. This requires that teachers have the adequate knowledge and skills to implement suitable activities, which are facilitated by accessible support for ECE centers and teachers. |
|  | Home-school cooperation | This relates to a two-way relationship between teachers and parents, such that activities in the ECE center may be followed up at home. This requires that both teachers and parents have the knowledge and understanding of the movement guidelines. |
|  | Community activities | This refers to the availability of activities in the community outside of school, which young children can join. This requires a mechanism for such information about activities to reach parents. |
| Challengers | Time poverty of parents | This refers to parents' lack of time to engage with their children to promote the movement guidelines, due to demanding work hours that also lead to fatigue and lack of energy. |
|  | Curriculum requirements | This refers to the traditional learning areas (e.g., literacy, numeracy) that are expected in the ECE curriculum. This effectively leads to teachers who perceive that the time for conducting movement activities is insufficient and who give significant amounts of homework. |
|  | Limited physical space | This observation applies to ECE classrooms, homes, and wider community resources (e.g., playgrounds, parks). This requires that activities be designed for small spaces. |
|  | Social values | This refers to beliefs that academic achievement is most important and that playing outdoors may have inherent associated risks (e.g., injuries, insect bites). |
|  | Pandemic-related restrictions | This refers to social distancing measures and school suspensions, which limited the spaces that children can use. The pandemic situation also diminished the contribution of the ECE centers in children's accrual of physical activity. |

## Discussion

Based on our survey findings, many ECE teachers in Hong Kong appear to have high awareness of the movement guidelines for young children, especially those with higher degree qualifications and longer working experiences. The majority of the teachers also claimed to have knowledge of the guidelines stipulate, but those who actually had the correct knowledge were much fewer. Parents, on the other hand, had limited awareness and knowledge of the movement guidelines. The domestic workers, who represent an extension of childcare at home, also reported limited awareness and knowledge of the movement guidelines. These findings suggest that dissemination strategies are crucially needed so that the adults who have influence on children’s movement behaviors would be aware, and have the correct knowledge of evidence-based recommendations.

In countries where movement guidelines have been disseminated, the stakeholders mainly included teachers and parents (14,15). The Hong Kong context introduced the roles played by additional carers (i.e., grandparents, domestic workers), which is comparable to other Asian cities and countries. The primary targets of our dissemination strategy remain the ECE teachers and the parents, who have control of the policies and activities in the classrooms and at home. They have also been known to be relevant agents that can influence the movement behaviours of children (26,27). While domestic workers look after the children during the parents’ working hours, our findings suggest that they do not have autonomy in how they deliver their roles in relation to the movement behaviors of children. There may be benefits to dissemination strategies for domestic workers, but such efforts should be through the parents given the dependency of domestic workers on the parents’ views. For example, we can provide parents with materials that they could readily share with domestic workers and grandparents (e.g., activity cards, YouTube videos).

There was a consensus across stakeholders and key informants that the movement guidelines adopted by the CHP are appropriate and suitable for young children in Hong Kong. However, reports from the stakeholders suggest that most children do not meet the guidelines, especially physical activity, while the limits of sedentary screen time and sleep recommendations seem to be achieved better. These findings are consistent with the reports from the Hong Kong health authorities (7). The likelihood of children meeting the guidelines point to the associated enablers and challenges that can be considered for designing dissemination strategies.

### Design of dissemination strategies

The factors that enable and challenge the uptake of the movement guidelines amongst young children in Hong Kong appear to cut across local (i.e., home, ECE center) and system contexts (i.e., curriculum policies, social expectations). Time is the most critical factor, which seems to impact on movement behaviors at home and in ECE centers. Time poverty, or having little time outside of work, has been identified as a critical barrier to an individual’s physical activity participation (28). In this study, we found how it manifests as a similar challenge in the context of parents and their children’s movement behaviors. For the ECE centers, time issues are related to the policies of the local education system, which affect how teachers allocate class time. ECE teachers in Hong Kong have noted that even the primary school curriculum has an impact on their priorities (29), because they need to ensure that children transition well from senior kindergarten. With the prevailing social values highlighted in this current study, movement behaviors tend to be less prioritized in ECE centers. Beliefs and attitudes of the wider society cannot be changed in the short-term; hence, dissemination of the movement guidelines will have to work around recognizing that academic pursuits will not be de-prioritized.

Dissemination strategies for teachers need to include components that will help them promote physical activities without taking time away from the academic learning areas. To this end, teachers can be supported to design learning activities where movement is integrated with academics (e.g., mathematics, language). It has been suggested that integrated activities have the potential to help children meet physical activity guidelines (30). In recent work in Hong Kong, integrated gross motor skills and mathematics successfully increased physical activity accrual of young children without any detrimental effect on mathematics abilities (31). Leveraging on the home-school cooperation, teachers can also design physically active homework for children.

Dissemination strategies for parents need to acknowledge the realities of working families and the impact of this on time. Like income, time is a finite resource, which parents need to allocate among competing demands (32). It is therefore necessary to help parents access a range of practical ideas for them to facilitate short bouts of physical activity at home. Online resources for physical activity, which had grown exponentially during the pandemic (33), can be curated and shared with parents (e.g., dancing or yoga can be done by children with little supervision required of parents). The use of such online resources also shifts the nature of screen time from being sedentary to active time.

The challenge of space limitation, which cuts across homes, schools, and community public spaces, is not surprising. High-rise housing is dominant in Hong Kong, where middle-class families live in relatively compact and small apartments (34) and public spaces tend to be small in mass housing areas compared to upmarket and commercial-business zones (35). Schools also have limited physical spaces, many of which have little opportunity to expand (36). In this context, teachers and parents tend to think that the lack of space prevents physical activities. However, physical activities can be taken up even in small spaces (37). Dissemination strategies need to provide teachers and parents with samples of activities that are designed for small spaces, including movements within compact households.

Not surprisingly, the impact of the COVID-19 pandemic was highly discussed by teachers and parents. Recent studies have shown decreased physical activity, increased sedentary behaviors, and disrupted sleep in children during the pandemic (38,39). The main restriction was related to the school suspension, where parents struggled with promoting the movement behaviors especially physical activity. In a post-pandemic context, our findings highlight the role of the ECE sector in promoting the movement guidelines. As the ECE teachers are key contributors to enhancing young children’s activity levels (40), dissemination strategies need to prepare them to intentionally provide opportunities that promote the uptake of the movement guidelines.

Bringing together the findings of this study, we recommend that in contexts where there are pervasive time poverty, space limitations, or high priority for academic achievements, dissemination strategies for the movement guidelines of young children should consider the following:

1. ensure that not only awareness, but also the correct knowledge of the guidelines is promoted among the adults that influence the movement behaviors of young children (i.e., teachers, parents, domestic workers, grandparents);
2. equip teachers with the ability to design and deliver intentional activities that integrate movement with academic learning areas, and that can be implemented in relatively small spaces;
3. direct parents to curated and publicly available resources which they can access for small-space home physical activities, managing sedentary screen time and sleep, and community activities that their children can join; such activities need to be viable despite parents having limited time after work;
4. enable the ECE centers to include practices that promote movement guidelines as they cooperate with the parents, and share information in their wider communities.

### Strengths and Limitations

Taking a broader approach to examining stakeholders’ views in relation to the movement guidelines for children, we adopted a mixed-methods design. This allowed us to assess the teachers’ awareness and knowledge levels across Hong Kong, thus setting us up for future monitoring. We did not do the same with parents, and future work is needed to enable a similar approach to monitoring. Grandparents were identified to have roles in caring for children, but we did not engage with them in this study. Further work is needed to explore their decision-making roles in relation to parents, and how movement guidelines are promoted to young children

While our findings are specifically drawn from the Hong Kong context, the enablers and challenges that we identified may be present in places where comparable circumstances exist (e.g., Macau, Singapore). In places where education policies place time constraints on ECE curricula, or the socio-economic environment leads to parents’ time poverty, dissemination of movement guidelines for children may consider the strategies identified through this research.

Our study was implemented during the COVID-19 pandemic. We gained insights that helped us better understand the movement behaviors of children in these unprecedented social circumstances. On the other hand, we also need to consider that the enablers and challenges that we identified may be in flux as we move forward in a post-pandemic world. As such, further work is mandatory so that we might be able to see long-term and sustainable promotion of healthy movement behaviors in young children. For Hong Kong, especially, future research is needed to address the social values that are viewed as challenges to physical activity in early childhood.

## Conclusion

We found that in Hong Kong, teachers are aware of the movement guidelines for young children, but their knowledge of the specific guidelines is inaccurate. On the other hand, parents and domestic workers have limited awareness and knowledge levels. We recommend that dissemination strategies in the ECE centers should not only deliver knowledge content, but also enable the stakeholders to mitigate the challenges associated with time and space limitations. Dissemination of similar movement guidelines in societies faced with similar conditions may consider adopting the recommendations that we presented. Finally, we acknowledge that true dissemination is an ongoing process that needs to involve stakeholders and policy-makers, for which future work is needed.

## Data Availability

All data produced in the present study are available upon reasonable request to the authors.

## Author Contributions

CMC is the principal investigator of the grant and is responsible for the overall concept and design of the study. RAJ, CSMN, CHPS, and KKHC are co-investigators of the grant and collaborated from grant preparation to project implementation. CMC drafted the manuscript, and all co-investigators provided expert input within their areas of expertise. All authors contributed to this paper and approve the submission.

## Acknowledgments

The investigators wish to thank the Centre for Health Protection of the Department of Health, Government of Hong Kong for the shared information on the movement guidelines for Hong Kong children.

